# A case-control and cohort study to determine the relationship between ethnic background and severe COVID-19

**DOI:** 10.1101/2020.07.08.20148965

**Authors:** Rosita Zakeri, Rebecca Bendayan, Mark Ashworth, Daniel M Bean, Hiten Dodhia, Stevo Durbaba, Kevin O’Gallagher, Claire Palmer, Vasa Curcin, Elizabeth Aitken, William Bernal, Richard D Barker, Sam Norton, Martin Gulliford, James TH Teo, James Galloway, Richard JB Dobson, Ajay M Shah

## Abstract

**Background:** People of minority ethnic background may be disproportionately affected by severe COVID-19 for reasons that are unclear. We sought to examine the relationship between ethnic background and (1) hospital admission for severe COVID-19; (2) in-hospital mortality.

**Methods:** We conducted a case-control study of 872 inner city adult residents admitted to hospital with confirmed COVID-19 (cases) and 3,488 matched controls randomly sampled from a primary healthcare database comprising 344,083 people resident in the same region. To examine in-hospital mortality, we conducted a cohort study of 1827 adults consecutively admitted with COVID-19. Data collected included hospital admission for COVID-19, demographics, comorbidities, in-hospital mortality. The primary exposure variable was self-defined ethnicity.

**Results:** The 872 cases comprised 48.1% Black, 33.7% White, 12.6% Mixed/Other and 5.6% Asian patients. In conditional logistic regression analyses, Black and Mixed/Other ethnicity were associated with higher admission risk than white (OR 3.12 [95% CI 2.63-3.71] and 2.97 [2.30-3.85] respectively). Adjustment for comorbidities and deprivation modestly attenuated the association (OR 2.28 [1.87-2.79] for Black, 2.66 [2.01-3.52] for Mixed/Other). Asian ethnicity was not associated with higher admission risk (OR 1.20 [0.86-1.66]). In the cohort study of 1827 patients, 455 (28.9%) died over a median (IQR) of 8 (4-16) days. Age and male sex, but not Black (adjusted HR 0.84 [0.63-1.11]) or Mixed/Other ethnicity (adjusted HR 0.69 [0.43-1.10]), were associated with in-hospital mortality. Asian ethnicity was associated with higher in-hospital mortality (adjusted HR 1.54 [0.98-2.41]).

**Conclusions:** Black and Mixed ethnicity are independently associated with greater admission risk with COVID-19 and may be risk factors for development of severe disease. Comorbidities and socioeconomic factors only partly account for this and additional ethnicity-related factors may play a large role. The impact of COVID-19 may be different in Asians.

**Funding sources:** British Heart Foundation (CH/1999001/11735 and RE/18/2/34213 to AMS); the National Institute for Health Research Biomedical Research Centre (NIHR BRC) at Guy’s & St Thomas’ NHS Foundation Trust and King’s College London (IS-BRC-1215-20006); and the NIHR BRC at South London and Maudsley NHS Foundation Trust and King’s College London (IS-BRC-1215-20018).

## Introduction

SARS-CoV2 (Severe acute respiratory syndrome coronavirus 2) is a highly transmissible respiratory pathogen that usually causes minor illness but in a small proportion of individuals leads to severe systemic disease (Coronavirus disease 2019 [COVID-19]) with 1-2% mortality.^1-4^ Older people, males, and those with comorbidities such as diabetes and cardiovascular disorders are over-represented among those requiring hospital admission.^1-4^ As COVID-19 spread to multi-ethnic populations in Western Europe and North America, numerous reports suggested that individuals of Black, Asian or Minority Ethnic background suffer a higher burden of severe disease.^5-9^ Audit data on patients admitted to UK intensive care units (ICU) with COVID-19 observed a substantially higher proportion of minority background patients than in previous years.^7^ US data reported higher per-capita mortality rates for African-American and Latino versus White people in several cities, though the reasons for these differences are unclear.^6^ A UK Office of National Statistics (ONS) analysis suggested that individuals of Black and South Asian descent had a higher likelihood of death than White people after adjustment for demographic and socioeconomic factors, but was limited by lack of information on comorbidities and the use of historic (2011) data for the reference population.^8^ The relative importance of contextual demographic and socioeconomic conditions and the prevalence of long-term morbidities remains unclear. These reports also do not distinguish between ethnicity-related differences in likelihood of infection, severity of initial disease or in-hospital survival.

COVID-19 mortality rates vary substantially by region and within urban conurbations. Nine of the ten UK local authorities with the highest age-standardised mortality rates are inner London Boroughs with high proportions of minority ethnic background people, socioeconomic deprivation and health inequalities.^10^ It is essential to dissect the complex relationship among ethnicity, socioeconomic factors and comorbidities to properly inform public health policy and mitigate the impact of COVID-19 on minority communities. We undertook an analysis of the relationship between ethnicity and severe COVID-19 in an inner London region with approximately 40% Black and minority ethnic background people among a population of 1.26 million. Our aims were to (1) determine the relationship between ethnicity, local population demography, socioeconomic profiles, individual-level comorbidities, and hospital admission for severe COVID-19; (2) establish whether ethnicity is associated with in-hospital outcome of severe COVID-19.

## Methods

### Study design and participants

We conducted an observational cohort study at King’s College Hospital Foundation Trust (KCHFT), which comprises two separate hospital sites in south London. We included consecutive adult patients (age ≥18 years) requiring emergency hospital admission with laboratory-confirmed COVID-19, between 1 March and 2 June, 2020. Eligible patients tested positive for viral RNA by quantitative RT-PCR in nasopharyngeal and oropharyngeal swabs (**Figure 1**).

**Figure 1.**
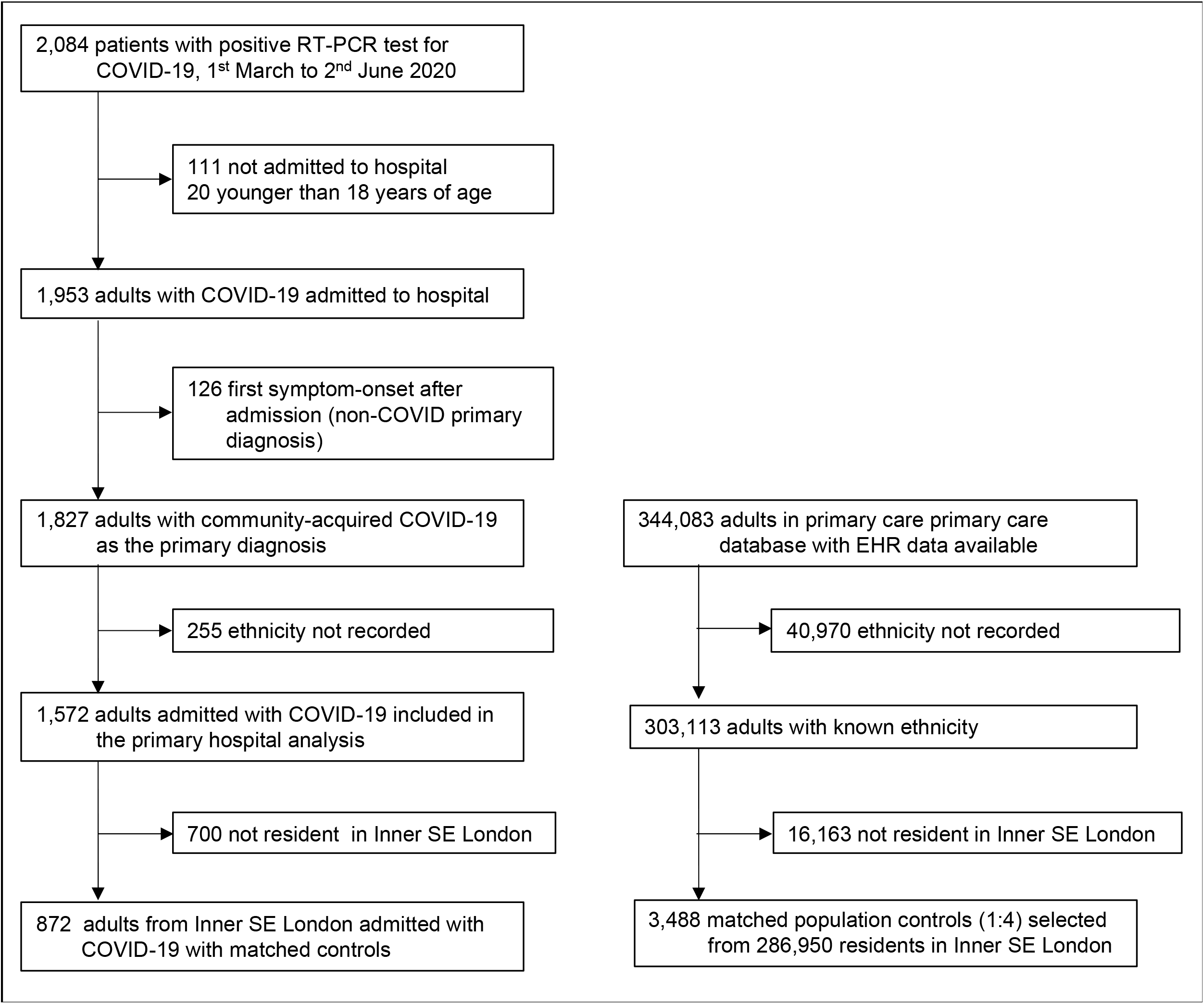
Study flow chart.

We performed a matched case-control (case-population) study with the subset of admitted patients who were inner city residents (>55% of the total hospital cohort). We matched COVID-19 patients (i.e. cases) with population controls sampled from the same inner city region using data from a primary healthcare database (Lambeth DataNet).^11,12^ This dataset includes de-identified data on 344,083 (96.8%) community-resident adults registered with 41 practices in inner south-east London.^13^ For each case, we randomly sampled four controls who were individually matched to cases by age (within 5-year age bands) and sex.

### Data sources and processing

Demographic and clinical data for admitted patients were retrieved from the electronic health record (EHR). We used well-validated natural language processing (NLP) informatics tools belonging to the CogStack ecosystem to access both structured fields and unstructured text in the EHR.^14-16^ Additional manual extraction, clinician case review, and mandatory hospital datasets were used for missing variables and validation. Individual-level anonymised primary care EHR data for the case-population study were extracted from the primary care database.

### Exposures

The primary exposure variable was self-reported ethnicity according to the 18 categories recommended by the ONS.^17^ These were reduced into four groups: White (British, Irish, Gypsy, any other White), Black (African, Caribbean, any other Black), Asian (Indian, Pakistani, Bangladeshi, Chinese, any other Asian), and Mixed/Other. Patients with missing ethnicity data were excluded. They tended to be younger and have fewer comorbidities in both the control population and admitted patients. Demographic and clinical variables, identified a priori as potential risk factors for severe COVID-19, included: age, sex, body mass index (BMI), smoking status, and comorbidities (asthma, chronic obstructive pulmonary disease [COPD], chronic kidney disease [CKD], coronary heart disease [CHD], diabetes, hypertension). For BMI, the most recent value within 6 months (median 27 days [LQ-UQ 0-38]) of admission (for hospital cases) or data extraction (primary care) was used. BMI categories were defined as underweight (<18.5 kg/m^2^), normal (18.5–24.9 kg/m^2^ [18.5-22.9 kg/m^2^ for Asians]), overweight (25–29.9 kg/m^2^ [23-27.4 kg/m^2^ for Asians]), and obese (≥30 kg/m^2^ [≥27.5 kg/m^2^ for Asians]).^18^ Comorbidities were categorised as present if recorded at any time in the EHR up to and including the day of admission (or data extraction in primary care). Multimorbidity was defined as 2 or more comorbidities.^19^ Socioeconomic status was estimated using the English Indices of Multiple Deprivation (IMD) score, presented in quintiles, as derived from residential postcode and Low Super Output Area (2019) codes.^20^ The same variable definitions were used across primary and secondary care, using internationally recognised terminology.

### Outcomes

Outcomes included hospital admission for COVID-19 and in-hospital mortality. The secondary outcome of ICU admission was evaluated in the hospital cohort. Start of follow-up for all analyses was taken as the date of self-reported symptom onset. Where symptom onset was unavailable (27%) and admission date was within 4 days of a COVID-positive RT-PCR test, admission date was used. This was based on the median duration between symptom onset and admission date for patients presenting with COVID-19. Outcomes were ascertained to 2 June 2020.

### Statistical analysis

Patient characteristics were summarised using frequency (%) and median with interquartile range (IQR). Comparisons across ethnic groups were made using the χ2 or Fisher’s exact test for categorical variables and 1-way ANOVA or Kruskal-Wallis test for continuous variables. Conditional logistic regression models were fitted to evaluate the association between ethnicity and hospital admission for COVID-19. All models were adjusted for the matching variables (age and sex) to eliminate residual confounding^21^ and based on their previously reported associations with COVID-19. Fully adjusted models included other potential confounding variables: IMD quintile and comorbidities as outlined above. White ethnicity was used as the reference group.

In the hospital cohort, we evaluated the association between ethnicity and risk of in-hospital death in a time-to-event framework using competing risks regression, based on Fine and Gray’s proportional sub-hazards model,^22^ with discharge from hospital assigned as a competing risk. For the secondary outcome of ICU admission, competing risks were death or discharge from hospital. Patient records were censored at the earlier of discharge from hospital or study end date. Univariable, age- and sex-adjusted, and fully adjusted multivariable models were performed as for the case-population study with White ethnicity as the reference group. Analyses were performed using STATA/IC (v16.1; StataCorp LLC, TX).

### Sensitivity analyses

Since BMI was missing in >30% of hospitalised patients, the primary analyses were performed without adjustment for BMI. To explore confounding by BMI, all analyses were repeated in the subset of patients with BMI data available. Additional sensitivity analyses were performed with imputation of missing variables using multiple imputations by chain equations.^23^

### Ethics

This study adhered to the principles of the UK Data Protection Act 2018 and UK National Health Service (NHS) information governance requirements. De-identified data from patients admitted to KCHFT were analysed under London SE Research Ethics Committee approval (reference 18/LO/2048) granted to the King’s Electronic Records Research Interface (KERRI). Specific work on COVID19 was approved by the KERRI committee which included patients and the Caldicott Guardian. Access to Lambeth DataNet was under a project-specific approval granted by Lambeth Public Health Caldicott Guardian, 30 April 2020; additional informed consent was not required.

## Results

### Relationship between COVID-19 admission and local demographic, health and socioeconomic profiles

Between March 1 and June 2, 2020, 1,827 adults were admitted with laboratory-confirmed COVID-19. 872 patients were inner city residents and had ethnicity data available (**Figure 1**). We compared this group with the population of adult residents registered with local primary care practices in the same region (n=286,950). COVID-19 patients were older (median age [IQR] 66 [55-80] versus 38 [29-51] years; p<0·001) and more likely to be male (55·9% versus 49·2%, p<0.001) than community residents, across all ethnic groups. However, there was a particularly high rate of admissions among Black people aged 45-65 years, as compared to the community age structure (**Supplemental Figure 1**). 65% of patients were from the lowest two quintiles of deprivation as compared to the 52% population prevalence.

To identify whether ethnicity was independently associated with hospital admission, each patient (n=872 cases) was matched by age and sex to randomly sampled population controls (n=3,488, **Figure 1**). Cases were more likely than controls to be of Black or Mixed/Other ethnicity and from the lowest deprivation quintile (**Supplemental Table 1**). Cases also had a greater prevalence of each of 6 comorbidities, overall (**Supplemental Table 1**) and within ethnic groups (**Supplemental Table 2**).

In analyses adjusting for the matching variables (age and sex) only, Black and Mixed/Other ethnicity were associated with higher odds of admission compared to White ethnicity (**Figure 2**) (OR [95% CI] for Black ethnicity 3·1 [2·6-3·7], Mixed/Other 3·0 [2·3-3·8]; both p<0·001). All comorbidities assessed were also associated with greater odds of admission, with the highest OR observed for hypertension (4·1 [3·4-4·9]) and diabetes (3·7 [3·1-4·3]). In fully adjusted models including comorbidities and deprivation quintile, there was modest attenuation of the association between Black or Mixed/Other ethnicity and risk of admission, with the OR still 2·3 to 2·7-fold higher for these groups (**Figure 2**). There was no increase in admission risk associated with Asian ethnicity although the proportion of Asian patients in our study was small (5.6% cases, 7.8% controls). Similar results were obtained in analyses adjusting for BMI, in the subset of individuals with BMI data available. BMI accounted for a small proportion of ethnicity-associated admission risk (**Supplemental Figure 2**).

**Figure 2.**
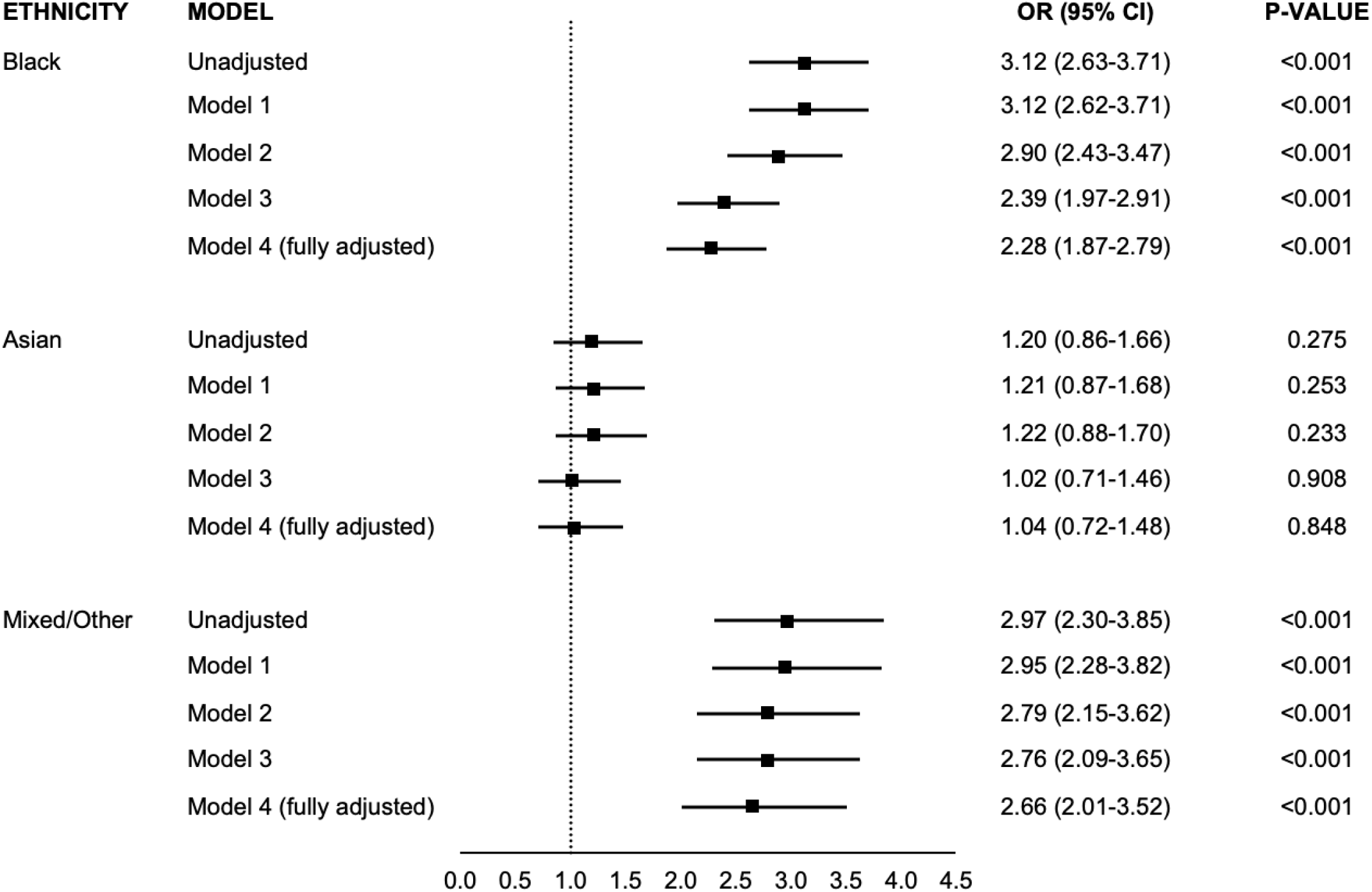
Association between ethnicity and risk of hospital admission for COVID-19. Odds ratios are compared to White ethnicity. Model 1 – adjusted for age and sex. Model 2 – adjusted for age, sex and index of multiple deprivation (IMD). Model 3 – adjusted for age, sex and comorbidities* Model 4 (fully adjusted model) – adjusted for age, sex, IMD, and comorbidities. *Comorbidities include asthma, chronic obstructive pulmonary disease, coronary heart disease, hypertension, diabetes, chronic kidney disease.

To assess whether the increased admission risk was specific for COVID-19, we examined all admissions from inner south-east London residents to the 5 main hospitals serving the region, both for COVID-19 (n=2434 patients; March 1 to June 2, 2020) and for respiratory infections over the preceding 12 months (n=7,081) (**Supplemental Figure 3**). These data confirmed a disproportionately higher number of minority ethnic group patients among those admitted for COVID-19 compared with emergency respiratory infection admissions in the preceding year.

### Clinical presentation of COVID-19 in patients requiring hospital admission

Among 1,827 patients admitted to KCHFT with COVID-19 from either inner city or suburbs, 1,572 had ethnicity data available (**Figure 1**). Patients of Black or minority ethnic background (46.3%) were significantly younger than White patients (median age [IQR]: 61 [51-76] versus 76 [63-86], p<0.001) but with a similar sex distribution (male sex: 56.4% BAME versus 56.3% White) (**Table 1**). Black patients had higher prevalence of hypertension, diabetes and CKD, and a higher proportion of overweight and obese individuals, than White patients. Conversely, Black patients had the lowest rates of CHD and COPD. Asian patients had higher prevalence of diabetes and obesity compared to White and a lower prevalence of COPD. 74% of Black patients resided in localities with the two lowest IMD quintiles as compared to 51% of Asian and 38% of White patients (p<0.001). Among patients with a recorded date for symptom onset (73.2%), the median duration of symptoms before admission was longer in minority ethnic groups (3 days for White, 4 days for Black, 7 days for Asian; p<0.001, **Table 2**) but did not vary substantially across IMD quintiles (median 4 days for both the most and least deprived quintiles).

**Table 1.** Baseline characteristics for patients admitted to hospital with COVID-19 by ethnic group

| CHARACTERISTIC | TOTAL<br>N=1572 | WHITE<br>N=845<br>(53.8%) | BLACK<br>N=486<br>(30.9%) | ASIAN<br>N=90<br>(5.7%) | MIXED / OTHER<br>N=151<br>(9.6%) | P-VALUE |
| --- | --- | --- | --- | --- | --- | --- |
| **Demographics** |  |  |  |  |  |  |
| Age, y | 70 (56-82) | 76 (63-86) | 61 (52-76) | 61 (44-78) | 60 (49-78) | <0.001 |
| Male sex | 886 (56.4) | 476 (56.3) | 269 (55.4) | 52 (57.8) | 89 (58.9) | 0.877 |
| BMI*, kg/m<sup>2</sup> | 26.7 (22.7-31.9) | 25.4 (22-30) | 28.6 (25-34) | 27.3 (25.6-30.7) | 27.5 (22.6-31.1) | <0.001 |
| Underweight | 66 (4.2) | 52 (6.2) | 6 (1.2) | 2 (2.2) | 6 (4.0) |  |
| Normal weight | 337 (21.4) | 241 (28.5) | 64 (13.2) | 8 (8.9) | 24 (15.9) |  |
| Overweight | 292 (18.6) | 159 (18.8) | 95 (19.5) | 15 (16.7) | 23 (15.2) |  |
| Obese | 340 (21.6) | 155 (18.3) | 130 (26.7) | 24 (26.7) | 31 (20.5) |  |
| Missing | 537 (34.2) | 238 (28.2) | 191 (39.3) | 41 (45.6) | 67 (44.3) |  |
| **Comorbidities** |  |  |  |  |  |  |
| Asthma | 193 (12.3) | 87 (10.3) | 70 (14.4) | 13 (14.4) | 23 (15.2) | 0.079 |
| COPD | 202 (12.9) | 152 (18.0) | 30 (6.2) | 6 (6.7) | 14 (9.3) | <0.001 |
| Coronary heart disease | 288 (18.3) | 187 (22.1) | 62 (12.8) | 17 (18.9) | 22 (18.3) | <0.001 |
| Hypertension | 1,027 (65.3) | 555 (65.7) | 348 (71.6) | 47 (52.2) | 77 (51.0) | <0.001 |
| Diabetes | 594 (37.8) | 242 (28.6) | 249 (51.2) | 42 (46.7) | 61 (40.4) | <0.001 |
| Chronic kidney disease | 405 (25.8) | 218 (25.8) | 146 (30.0) | 17 (18.9) | 24 (15.9) | 0.002 |
| Number of chronic conditions<br>(excluding obesity) |  |  |  |  |  | <0.001 |
| None | 346 (22.0) | 190 (22.5) | 81 (16.7) | 23 (25.6) | 52 (34.4) |  |
| 1 | 376 (23.9) | 207 (24.5) | 115 (23.7) | 24 (26.7) | 30 (19.9) |  |
| 2+ | 850 (54.1) | 448 (53.0) | 290 (59.7) | 43 (47.8) | 69 (45.7) |  |
| **Socioeconomic factors** |  |  |  |  |  |  |
| Deprivation quintile |  |  |  |  |  | <0.001 |
| 1 (most deprived) | 331 (24.2) | 117 (13.8) | 161 (33.1) | 17 (18.9) | 36 (23.8) |  |
| 2 | 487 (31.0) | 200 (23.7) | 199 (40.9) | 29 (32.2) | 59 (39.1) |  |
| 3 | 280 (17.8) | 140 (16.6) | 89 (18.3) | 17 (18.9) | 34 (22.5) |  |
| 4 | 239 (15.2) | 195 (23.1) | 21 (4.3) | 10 (11.1) | 13 (8.6) |  |
| 5 (least deprived) | 225 (14.3) | 189 (22.4) | 12 (2.5) | 17 (18.9) | 7 (4.6) |  |
| Missing | 10 (0.6) | 4 (0.4) | 4 (0.8) | - | 2 (1.3) |  |

Data are presented as number (%) or median (IQR).
\* BMI was categorised as underweight (<18.5 kg/m<sup>2</sup>), normal (18.5–24.9 kg/m<sup>2</sup> [18.5–22.9 kg/m<sup>2</sup> for Asians]), overweight (25–29.9 kg/m<sup>2</sup> [23–27.4 kg/m<sup>2</sup> for Asians]), and obese (≥30 kg/m<sup>2</sup> [≥27.5 kg/m<sup>2</sup> for Asians]).

**Table 2.**
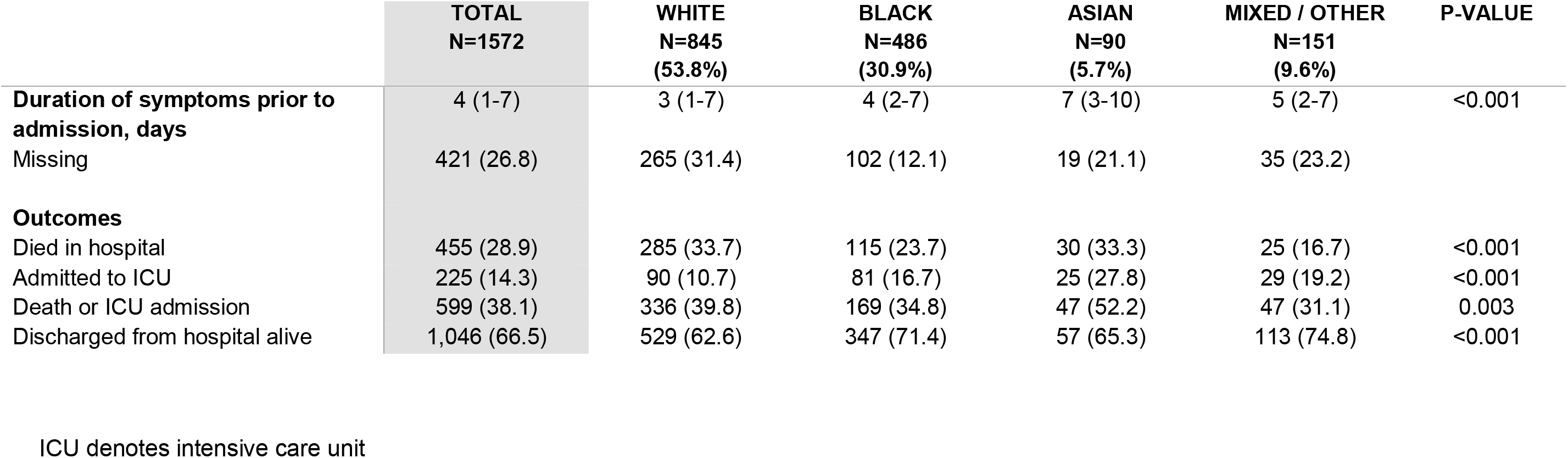
Duration of symptoms before admission and in-hospital outcomes for patients admitted with COVID-19 by ethnic group

### Risk of in-hospital mortality with COVID-19

By 2 June 2020, 455 of 1,572 patients (28.9%) had died and 1,046 (66.5%) had been discharged. The median (IQR) length of stay was 8 (4-16) days. Age-adjusted cumulative incidence curves for in-hospital mortality by ethnic group are displayed in **Figure 3A** and stratified by sex in **Supplemental Figure 4**.

**Figure 3.**
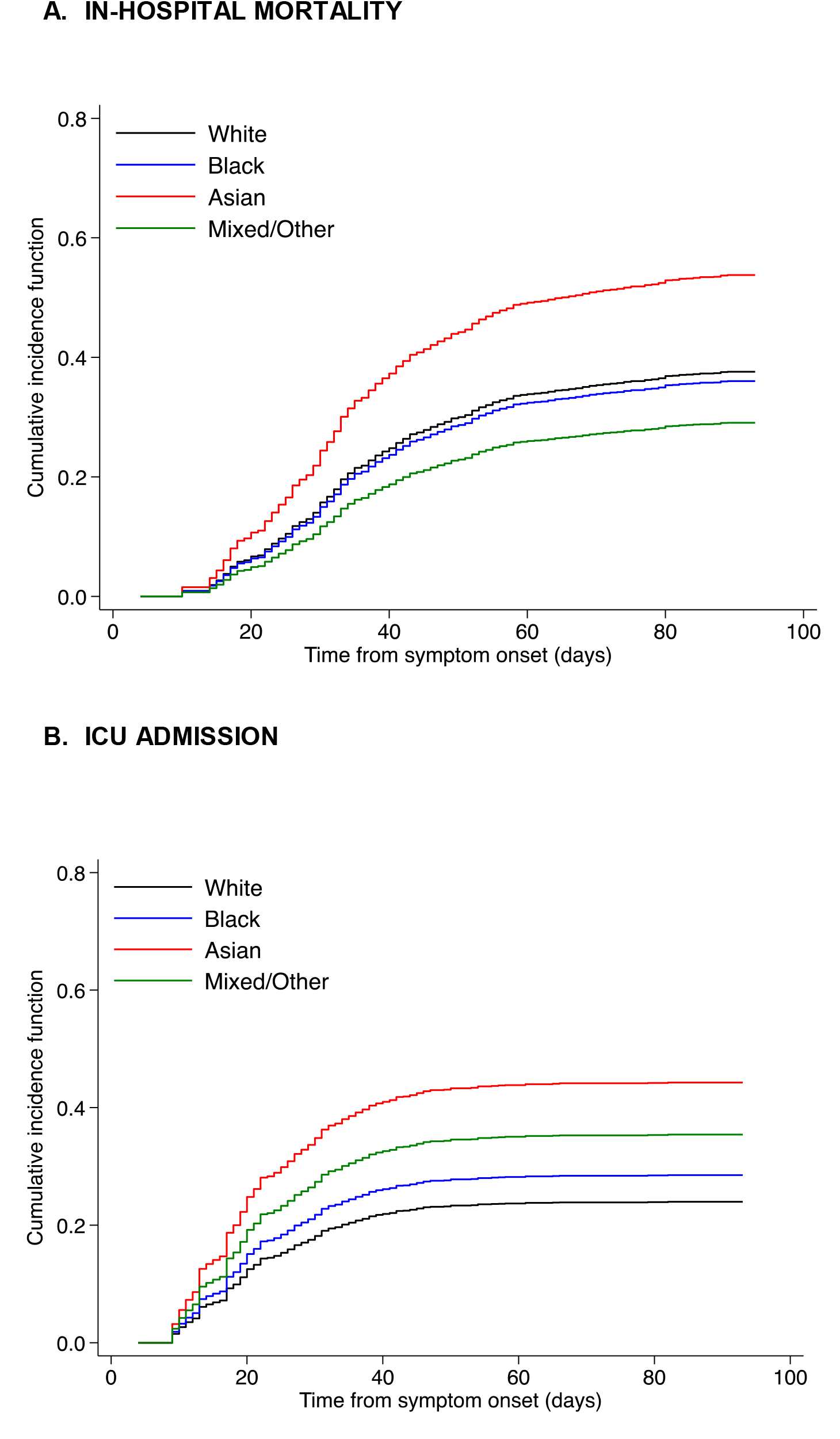
Cumulative incidence of in-hospital mortality or ICU admission by ethnicity, adjusted for age.

In unadjusted analyses, Black or Mixed/Other ethnicity were associated with a lower risk of death compared to White, however this was largely due to their younger age (**Figure 4**). In age- and sex-adjusted and full adjusted analyses, the risk of death was not significantly different between Black or Mixed/Other ethnicity versus White. The adjusted analyses estimated increased mortality risk with Asian ethnicity but with considerable uncertainty; the number of Asian patients was relatively low in our cohort.

**Figure 4.**
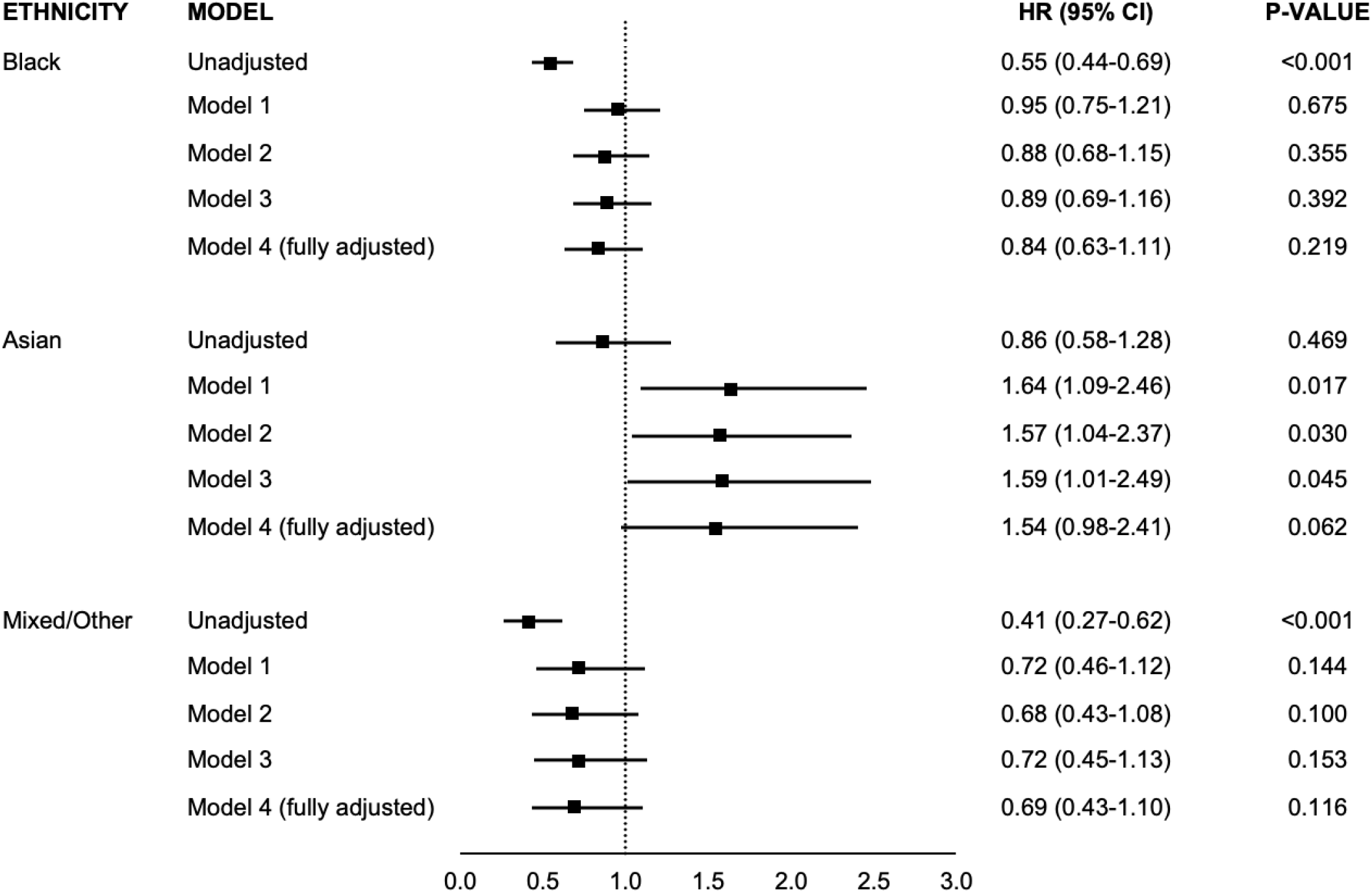
Association between ethnicity and risk of in-hospital mortality with COVID-19. Hazard ratios are compared to White ethnicity. Model 1 – adjusted for age and sex. Model 2 – adjusted for age, sex and index of multiple deprivation (IMD). Model 3 – adjusted for age, sex and comorbidities* Model 4 (fully adjusted model) – adjusted for age, sex, IMD, and comorbidities. *Comorbidities include asthma, chronic obstructive pulmonary disease, coronary heart disease, hypertension, diabetes, chronic kidney disease.

Age-adjusted cumulative incidence curves for admission to ICU by ethnic group, and stratified by sex, are displayed in **Figure 3B** and **Supplemental Figure 5**, respectively. Asian patients had significantly higher rates of ICU admission even after adjustment for age, sex, comorbidities and IMD (**Figure 5**).

**Figure 5.**
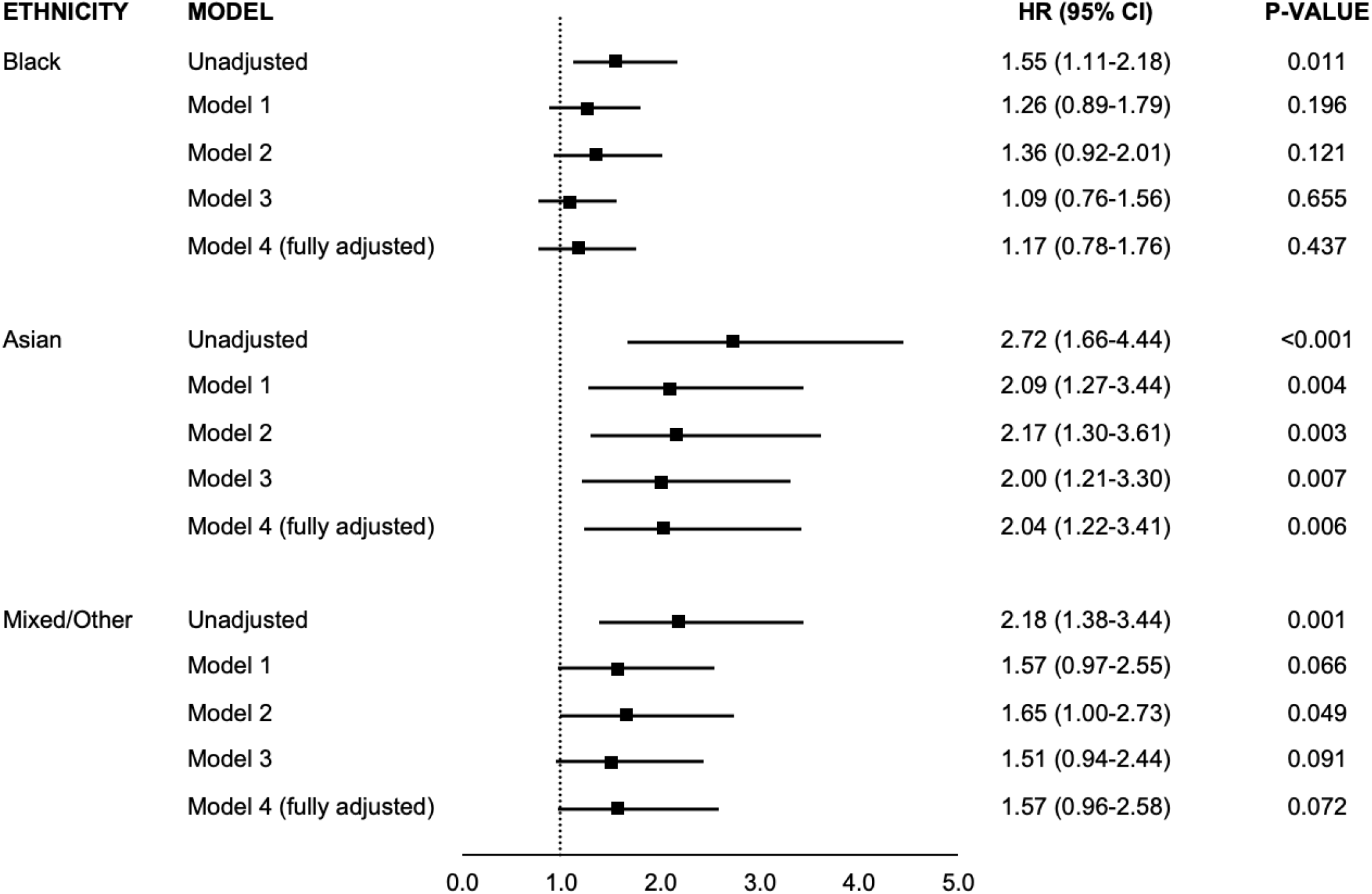
Association between ethnicity and risk of ICU admission with COVID-19. Hazard ratios are compared to White ethnicity Model 1 – adjusted for age and sex Model 2 – adjusted for age, sex and index of multiple deprivation (IMD) Model 3 – adjusted for age, sex and comorbidities* Model 4 (fully adjusted model) – adjusted for age, sex, IMD, and comorbidities *Comorbidities include asthma, chronic obstructive pulmonary disease, coronary heart disease, hypertension, diabetes, chronic kidney disease.

### Sensitivity analyses

Sensitivity analyses adjusting for BMI in the subset of patients with BMI available produced results similar to those in the main analysis (**Supplemental Figure 6**). The association between ethnicity and in-hospital mortality remained consistent across age strata (**Supplemental Figure 7**). Finally, using multiple imputation, fully adjusted hazard ratios for in-hospital mortality remained similar to the main analysis.

## Discussion

People of Black and Minority ethnic background are reported to have a disproportionately high mortality from COVID-19 both in the US and the UK, but it is not established whether this is due to increased risk of infection, greater frequency of severe disease once infected, or worse in-hospital survival.^5-9^ The relative importance of socioeconomic deprivation, health inequalities or other underlying factors is also unclear. The impact of COVID-19 on minority communities is especially high in inner cities which have a multi-ethnic population.

The current study makes several new findings. First, we employ a case-population study to identify an approximately 3-fold higher risk of hospital admission with COVID-19 for Black and Mixed ethnicity groups (but not Asians) compared to White among inner city residents. After adjusting for comorbidities and socioeconomic deprivation score, there remains a 2·3 to 2·7-fold higher admission risk.

Secondly, we find inter-ethnic variation in demographics and clinical characteristics in admitted patients. Minority ethnic group patients are on average 10-15 years younger than White patients yet have a higher prevalence of chronic medical conditions. Diabetes is especially prevalent in all non-white groups while Black patients also have high rates of hypertension and CKD.

Thirdly, we find no association between Black or Mixed ethnicity and in-hospital outcome. An association with increased in-hospital mortality is found for Asian patients but should be cautiously interpreted due to the low numbers in this group. Overall, our results suggest that the higher COVID-19 mortality reported for Black people in prior studies is most likely driven by a higher rate of admissions rather than a worse in-hospital outcome.

Disproportionately higher admission of Black and Mixed ethnicity individuals compared to White people living in the same region may be driven either by an increased risk of infection or more aggressive early disease progression or both. An increased susceptibility to infection could occur for several reasons. It has been suggested that a higher prevalence of socioeconomic deprivation (e.g. with poor housing) and cultural factors such as living in multi-generational households may increase susceptibility in some ethnic groups.^5,24^ Adjustment for deprivation as assessed by IMD quintile had only a modest effect in our study. However, it is recognised that the complexity of disadvantage related to socioeconomic factors may be incompletely captured by the IMD score and that area-based metrics may not be directly comparable across ethnic groups.^25^ Minority ethnic groups in the US and Europe have a higher prevalence of comorbidities such as diabetes compared with White groups,^26, 27^ which may also increase susceptibility to infection and seemed to contribute to part of the additional admission risk. Beyond higher prevalence, the adverse effects of some long-term conditions are disproportionately amplified by specific ethnicity. For example, diabetes is more likely to progress to complications in African Americans, African Caribbeans and South Asians compared to White individuals.^27,28^ Similarly, the likelihood of incident cardiovascular disease in people with metabolic risk factors is amplified by African Caribbean and South Asian ethnicity.^29^ Therefore, ethnicity-related amplification of comorbidity risks as well as latent socioeconomic factors could potentially contribute to part of the comorbidity- and deprivation-independent risk of admission for COVID-19.

An alternative driver of higher admission risk in Black and Mixed ethnicity people may be that they are more likely once infected to progress to disease that requires admission (noting that an increase in mild infections per se should not affect admission risk or mortality). It is notable that the ethnicity distribution for admissions with respiratory infections in the same community population was markedly different from that for COVID-19 admissions, suggesting that more complex factors are involved with COVID-19. A higher prevalence of comorbidities such as diabetes could potentially contribute to more frequent progression to severe disease given that it enhances pro-inflammatory effects. Previous studies have shown that immune responses to pathogens are significantly different between individuals of African versus European ancestry, with African ancestry associated with a stronger inflammatory response.^30,31^ Our study was, however, not designed to distinguish between infection risk and more severe early disease as drivers of COVID-19 admission.

We did not find evidence that in-hospital survival was significantly different between Black or Mixed ethnicity and White patients, suggesting that there are no major differences between these groups in life-threatening complications. The age-adjusted incidence of ICU admission was also similar for Black and White patients. A strong trend towards higher in-hospital death and admission to ICU was noted in Asians compared to the other groups. We also observed the longest symptom duration prior to admission in Asians, followed by Black and Mixed ethnicity patients, and then White patients. Whether this reflects difficulties in access to healthcare, differences in health-seeking behaviours, or is related to disease severity and progression requires further investigation. The in-hospital mortality data for Asians also need to be further explored in studies with larger numbers of Asian patients as well as for the different Asian sub-categories.

Most previous studies that addressed the relationship between ethnicity and COVID-19 either focused solely on mortality referenced to aggregate populations or made causal inferences from investigation of hospitalised patients.^7-9^ The limitations of these approaches in terms of selection or collider bias have been discussed.^32^ In our study, the use of a case-population design to assess the risk of admission for severe COVID-19 allowed us to compare the characteristics of admitted patients with a representative sample of the source population and thereby minimise selection bias. Our primary care database covered a large inner city region that closely matched the normal catchment area for the hospital. We cannot exclude the possibility that White patients are differentially admitted to other hospitals in the area but we consider this unlikely since emergency admissions are typically to the nearest hospital in the UK National Health Service. Moreover, we present data for all five hospitals in south-east London, which confirm excess COVID-19 admissions in the Black ethnic group. It is unlikely that our findings are explained by a lower threshold for admission in the Black group, because symptom duration was generally greater for these patients. This approach strengthens our conclusions regarding the interrelationship between ethnicity, comorbidities, deprivation score and risk of admission. Although our study was performed in a single region of inner London, the results are likely to be applicable to similar large cities with multi-ethnic populations around the world, especially in relation to the risks that appear independent of socioeconomic deprivation.

Our study has some limitations. In common with the majority of published studies so far, we did not have data on the exposure to SARS-CoV2 in the community nor on the prevalence of asymptomatic or mild disease. Our conclusions regarding risk of infection are therefore limited by this unknown factor. A small proportion of our cohort had missing ethnicity, consistent with other similar studies. While information on deprivation related to locality was available, we did not have individual-level data on factors such as number of people in the household, occupation, availability of personal protective material, and adherence to social distancing measures. Detailed information on the degree of control of comorbidities was also unavailable. Conclusions regarding obesity were limited by the high proportion of patients of all ethnic groups for whom data on BMI was unavailable. However, we performed sensitivity analyses to exclude the possibility that our conclusions were confounded by these missing data. Finally, the ethnic categories used in our study were broad categories and the analyses do not explore variation within an ethnic category, e.g. Black African versus Black Caribbean. Despite these limitations, the current findings on risk of admission related to Black and Mixed ethnicity and the lack of association with in-hospital outcome for the individual patient provide new insights to inform public health measures to tackle risk.

## Conclusions

Our results indicate that increased admission risk rather than worse in-hospital outcome may be the main contributor to higher mortality in Black patients. They also suggest that beyond the higher prevalence of comorbidities (especially diabetes and hypertension) and the socioeconomic conditions captured by IMD, additional ethnicity-related factors contribute to the association with admission for COVID-19. The identification of these factors, whether latent socioeconomic or biological, is an important area for continuing research with potentially global relevance. In the meantime, ethnic background may be considered a risk factor for susceptibility to severe COVID-19, in addition to factors such as age, male sex and the presence of comorbidities.

## Data Availability

The authors declare that all data supporting the findings of this study are available within the
article and its supplementary information files.

## Author contributions

JTHT, JG, RJBD and AMS conceived the study. RZ, MA, DMB, SN and MG participated in the study design. RZ, HD, SD, KOG, CP, VC, EA, WB, RDB, JTHT and JG participated in the data collection. RZ, RB, DMB and SN did data analyses. RZ, RB, MA, WB, RDB, SN, MG, JTHT, JG, RJDB and AMS contributed to data interpretation. RZ and AMS drafted the first version of the manuscript. All authors contributed to and approved the final manuscript and the decision to submit. The corresponding author attests that all listed authors meet authorship criteria and that no others meeting the criteria have been omitted.

## Availability of data and materials

The authors declare that all data supporting the findings of this study are available within the article and its supplementary information files.

## Acknowledgements

We are extremely grateful to all the clinicians and other staff who have looked after our patients. We thank the patient experts of the KERRI committee and Professor Clive Kay, Professor Alastair Baker, Professor Irene Higginson and Professor Jules Wendon for their support. We gratefully acknowledge the excellent help of Chris Fry, Dominic Thurgood and Isuf Ali in the KCHFT Business Intelligence Unit.

## SUPPLEMENTAL MATERIALS

**Supplementary Table 1.**
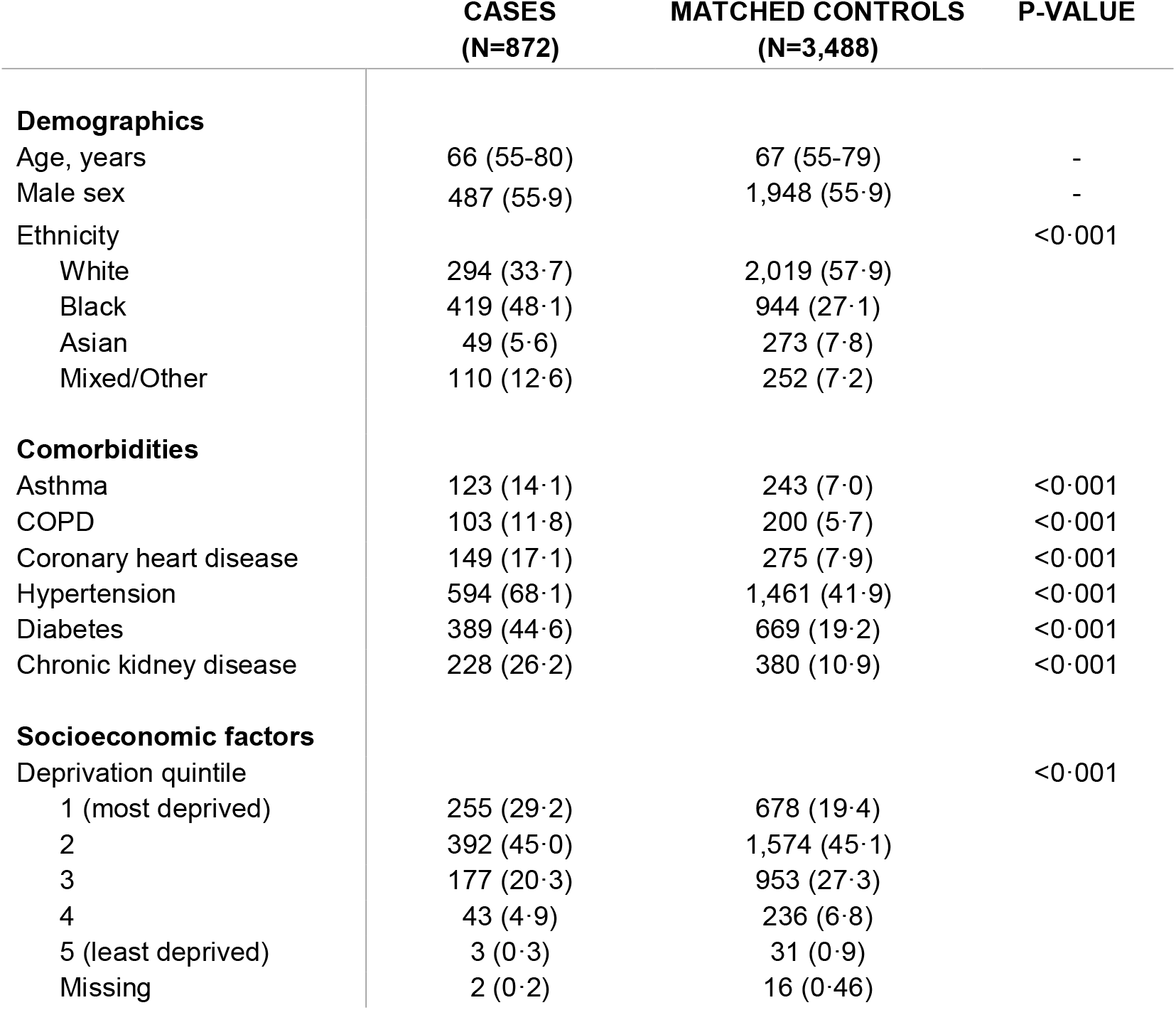
Characteristics of cases and population controls

|  | **CASES**<br>**(N=872)** | **MATCHED CONTROLS**<br>**(N=3,488)** | **P-VALUE** |
| --- | --- | --- | --- |
| **Demographics** |  |  |  |
| Age, years | 66 (55-80) | 67 (55-79) | - |
| Male sex | 487 (55.9) | 1,948 (55.9) | - |
| Ethnicity |  |  | <0.001 |
| White | 294 (33.7) | 2,019 (57.9) |  |
| Black | 419 (48.1) | 944 (27.1) |  |
| Asian | 49 (5.6) | 273 (7.8) |  |
| Mixed/Other | 110 (12.6) | 252 (7.2) |  |
| **Comorbidities** |  |  |  |
| Asthma | 123 (14.1) | 243 (7.0) | <0.001 |
| COPD | 103 (11.8) | 200 (5.7) | <0.001 |
| Coronary heart disease | 149 (17.1) | 275 (7.9) | <0.001 |
| Hypertension | 594 (68.1) | 1,461 (41.9) | <0.001 |
| Diabetes | 389 (44.6) | 669 (19.2) | <0.001 |
| Chronic kidney disease | 228 (26.2) | 380 (10.9) | <0.001 |
| **Socioeconomic factors** |  |  |  |
| Deprivation quintile |  |  | <0.001 |
| 1 (most deprived) | 255 (29.2) | 678 (19.4) |  |
| 2 | 392 (45.0) | 1,574 (45.1) |  |
| 3 | 177 (20.3) | 953 (27.3) |  |
| 4 | 43 (4.9) | 236 (6.8) |  |
| 5 (least deprived) | 3 (0.3) | 31 (0.9) |  |
| Missing | 2 (0.2) | 16 (0.46) |  |

**Supplementary Table 2.** Characteristics of cases and age-sex-matched population controls stratified by ethnicity

|  | WHITE |  | BLACK |  | ASIAN |  | MIXED/OTHER |  |
| --- | --- | --- | --- | --- | --- | --- | --- | --- |
|  | Cases<br>N=294 | Controls<br>N=2,019 | Cases<br>N=419 | Controls<br>N=944 | Cases<br>N=49 | Controls<br>N=273 | Cases<br>N=110 | Controls<br>N=252 |
| **Comorbidities** |  |  |  |  |  |  |  |  |
| Asthma | 34 (11.6) | 129 (6.4) | 67 (16.0) | 68 (7.2) | 7 (14.3) | 28 (10.3) | 15 (13.6) | 18 (7.1) |
| COPD | 62 (21.1) | 152 (7.5) | 27 (6.4) | 30 (3.2) | 4 (8.2) | 10 (3.7) | 10 (9.1) | 8 (3.2) |
| Coronary heart disease | 64 (21.8) | 174 (8.6) | 57 (13.6) | 60 (6.4) | 9 (18.4) | 29 (10.6) | 19 (17.3) | 12 (4.8) |
| Hypertension | 200 (68.0) | 714 (35.4) | 305 (72.8) | 537 (56.9) | 32 (65.3) | 120 (44.0) | 57 (51.8) | 90 (35.7) |
| Diabetes | 97 (33.0) | 255 (12.6) | 222 (53.0) | 275 (29.1) | 27 (55.1) | 91 (33.3) | 43 (39.1) | 48 (19.1) |
| Chronic kidney disease | 68 (23.1) | 202 (10.0) | 129 (30.8) | 128 (13.6) | 13 (26.5) | 33 (12.1) | 18 (16.4) | 17 (6.8) |
| **Multimorbidity\*** | 161 (54.8) | 441 (21.8) | 259 (61.8) | 328 (34.8) | 30 (61.2) | 98 (35.9) | 52 (47.3) | 55 (21.8) |
| **BMI** |  |  |  |  |  |  |  |  |
| Underweight | 19 (6.5) | 63 (3.1) | 6 (1.4) | 16 (1.7) | 1 (2.0) | 11 (4.0) | 3 (2.7) | 6 (2.4) |
| Normal weight | 76 (25.9) | 789 (39.1) | 54 (12.9) | 223 (23.6) | 3 (6.1) | 51 (18.7) | 17 (15.5) | 73 (29.0) |
| Overweight | 60 (20.4) | 648 (32.1) | 82 (19.6) | 340 (36.0) | 8 (16.3) | 112 (41.0) | 17 (15.5) | 79 (31.3) |
| Obese | 51 (17.3) | 396 (19.6) | 116 (27.7) | 322 (34.1) | 14 (28.6) | 77 (28.2) | 22 (20.0) | 64 (25.4) |
| Missing | 88 (29.9) | 123 (6.1) | 161 (38.4) | 43 (4.6) | 23 (46.9) | 22 (8.1) | 51 (46.4) | 30 (11.9) |
| **Socioeconomic factors** |  |  |  |  |  |  |  |  |
| IMD quintile |  |  |  |  |  |  |  |  |
| 1 (most deprived) | 74 (25.2) | 302 (15.0) | 139 (33.2) | 278 (29.4) | 12 (24.5) | 34 (12.5) | 30 (27.3) | 64 (25.4) |
| 2 | 127 (43.2) | 885 (43.8) | 186 (44.4) | 469 (49.7) | 26 (53.1) | 105 (38.5) | 53 (48.2) | 115 (45.6) |
| 3 | 64 (21.8) | 631 (31.3) | 80 (19.1) | 164 (17.4) | 10 (20.4) | 98 (35.9) | 23 (20.9) | 60 (23.8) |
| 4 | 27 (9.2) | 161 (8.0) | 11 (2.6) | 28 (3.0) | 1 (2.0) | 34 (12.5) | 4 (3.6) | 13 (5.2) |
| 5 (least deprived) | 1 (0.3) | 27 (1.3) | 2 (0.5) | 2 (0.2) | 0 | 2 (0.7) | 0 | 0 |
| Missing | 1 (0.3) | 13 (0.6) | 1 (0.2) | 3 (0.3) | - | - | - | - |

BMI, body mass index; COPD, chronic obstructive pulmonary disease; IMD, index of multiple deprivation
\* Defined as $\geq 2$ comorbidities

**Supplementary Figure 1.**
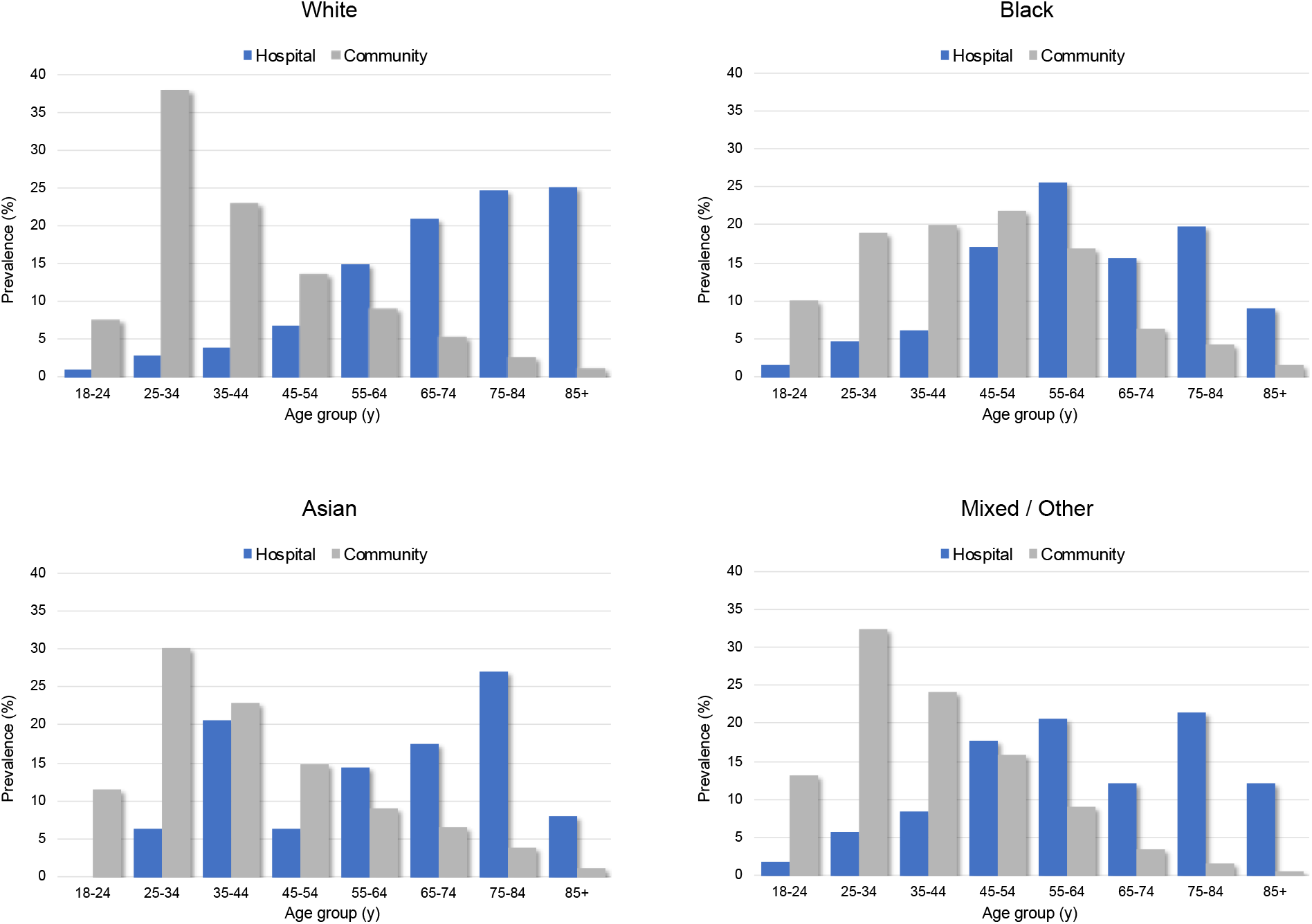
Age distribution of patients admitted to hospital with COVID-19 from Inner South East London and community residents by ethnic group. The prevalence is the percentage of individuals in the relevant age band for that ethnic group (e.g. 38% of all White adults in the community are aged 25-34).

**Supplementary Figure 2.**
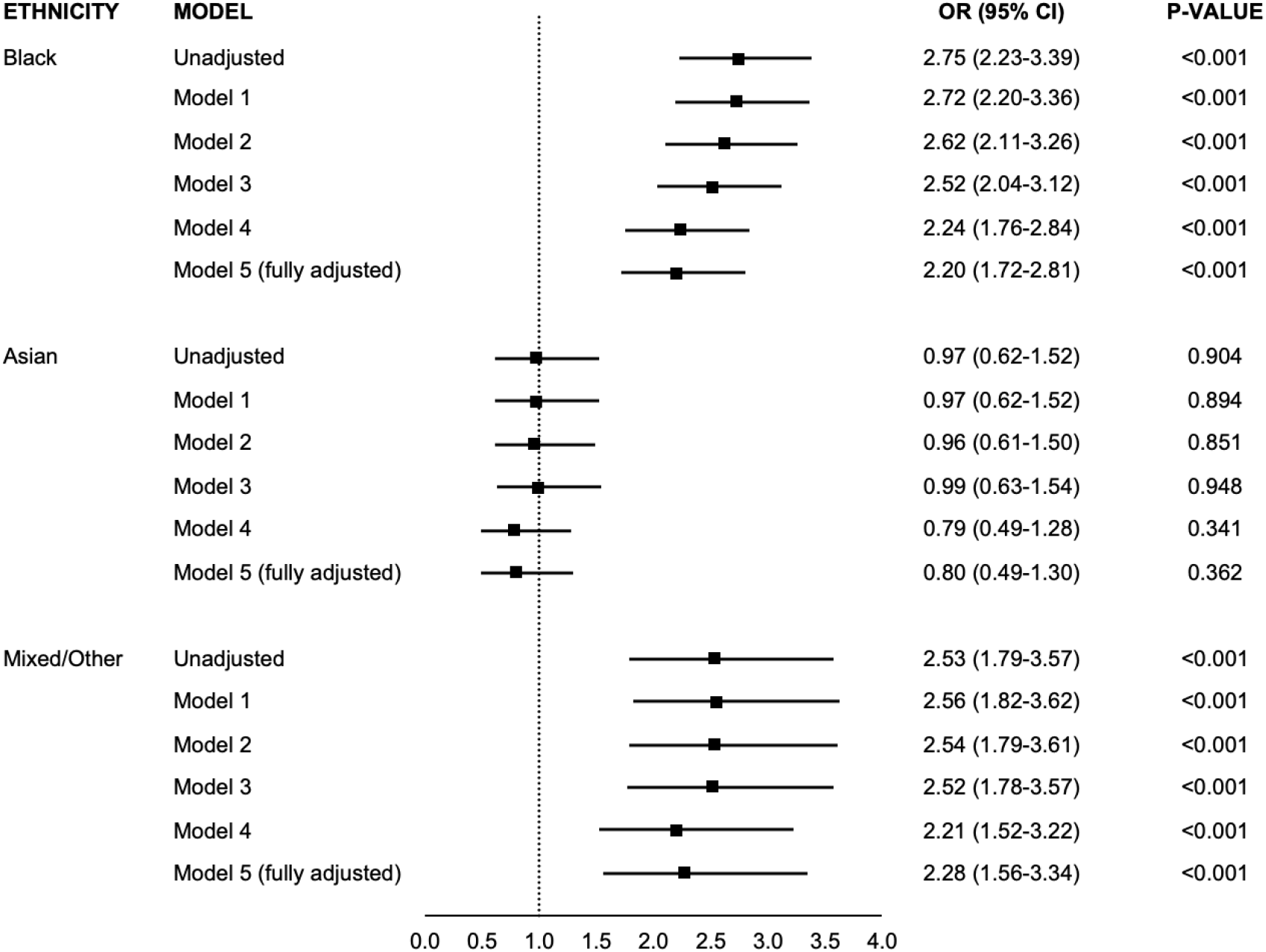
Association between ethnicity and risk of COVID-19 admission in the subset of patients with BMI data available. Odds ratios are compared to White ethnicity Model 1 – adjusted for age and sex Model 2 – adjusted for age, sex and index of multiple deprivation (IMD) Model 3 – adjusted for age, sex and body mass index (BMI) Model 4 – adjusted for age, sex and comorbidities* Model 5 (fully adjusted model) – adjusted for age, sex, IMD, BMI, and comorbidities *Comorbidities include asthma, chronic obstructive pulmonary disease, coronary heart disease, hypertension, diabetes, chronic kidney disease.

**Supplementary Figure 3.**
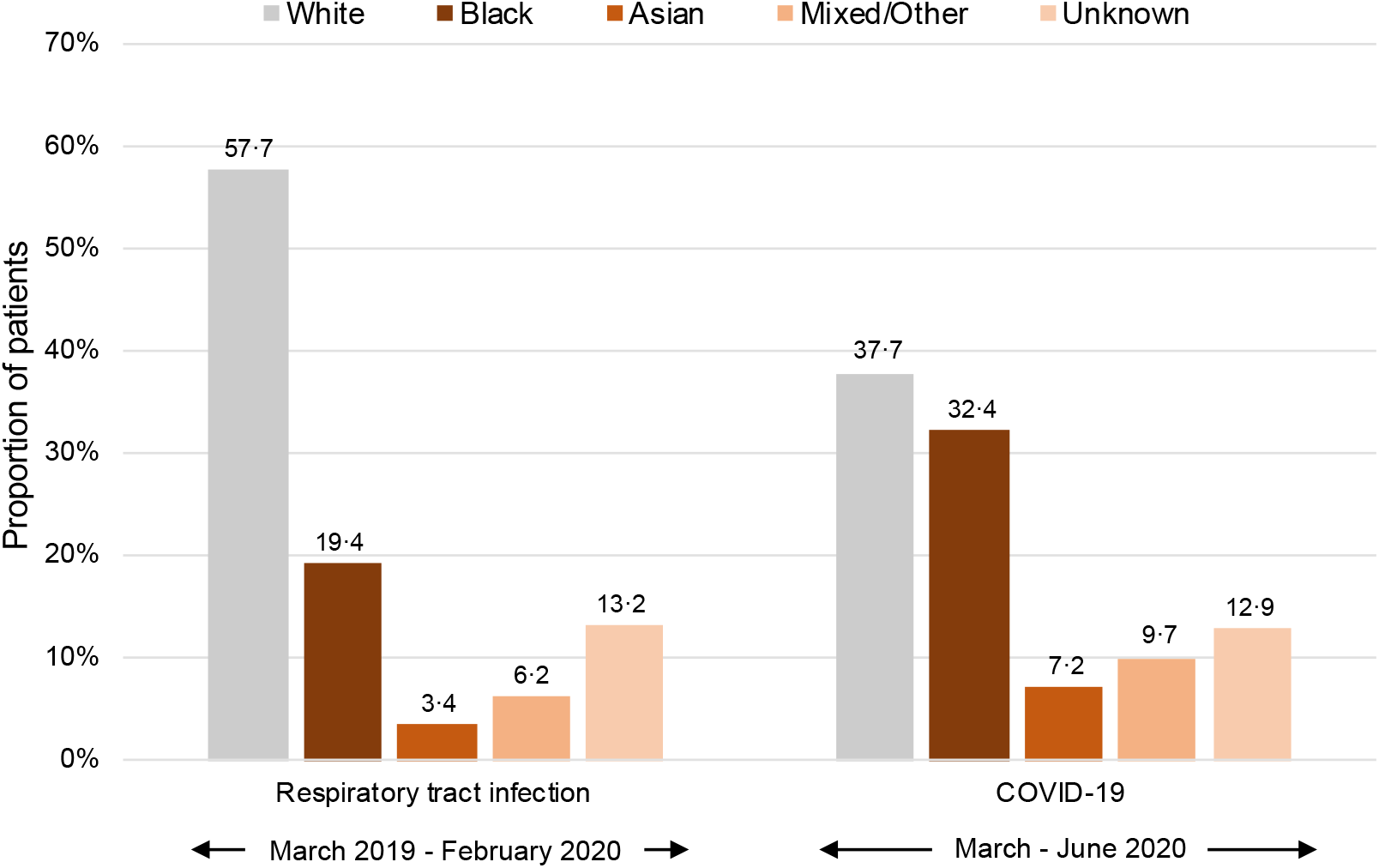
Ethnicity distribution of emergency hospital admissions in Inner South East London. Data on pre-COVID-19 era (1 March 2019 to 29 February 2020) emergency admissions to hospitals within inner SE London (namely KCHFT Denmark Hill, Guy’s & St Thomas’ NHS Foundation Trust, and Lewisham & Greenwich NHS Trust) were retrieved from the Healthcare Evaluation Data system (HED; www.hed.nhs.uk) on 2 June 2020. HED facilitates analyses of NHS Digital’s Hospital Episode Statistics (HES) and ONS datasets. Anonymised data on numbers of admissions with COVID-19 and ethnicity breakdown were obtained directly from these three NHS Trusts. These data represent 90-95% of all emergency admission from patients who are resident in this region.

**Supplementary Figure 4.**
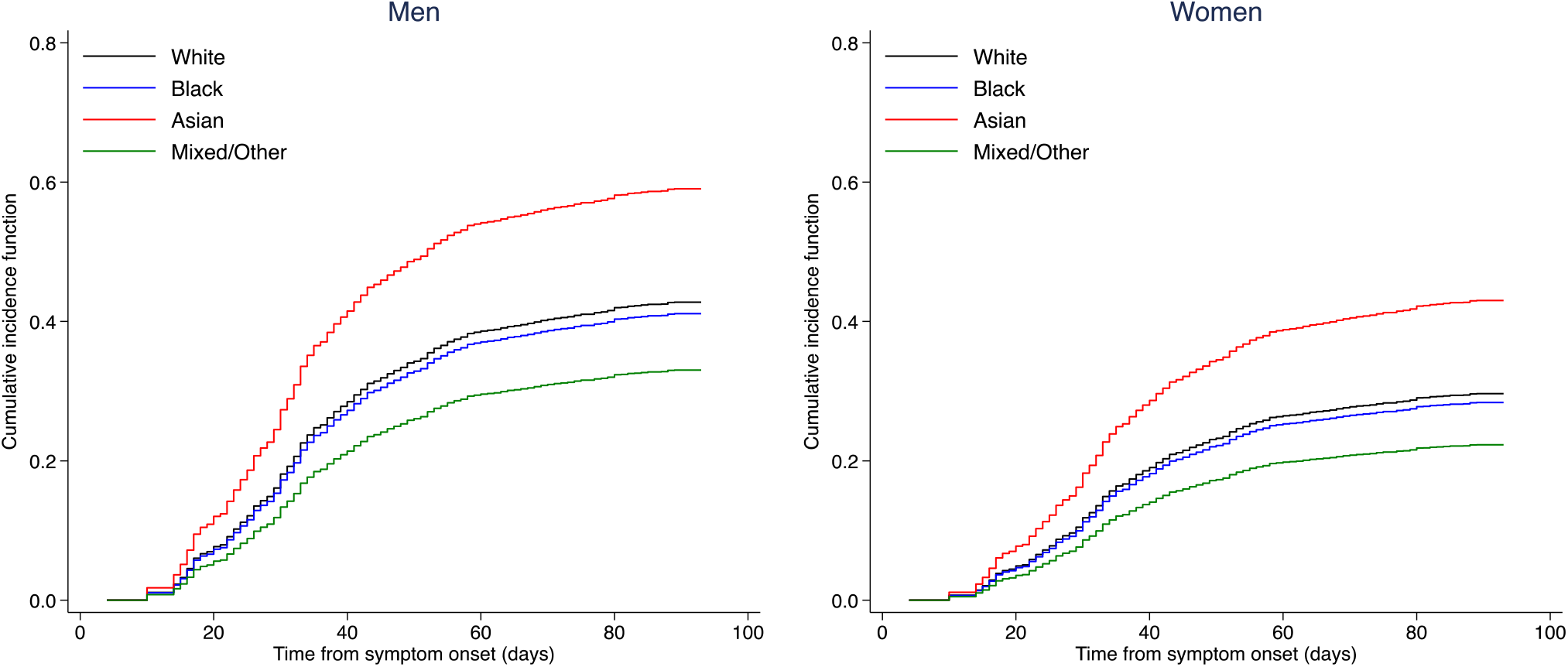
Cumulative age-adjusted incidence of in-hospital mortality by sex and ethnicity

**Supplementary Figure 5.**
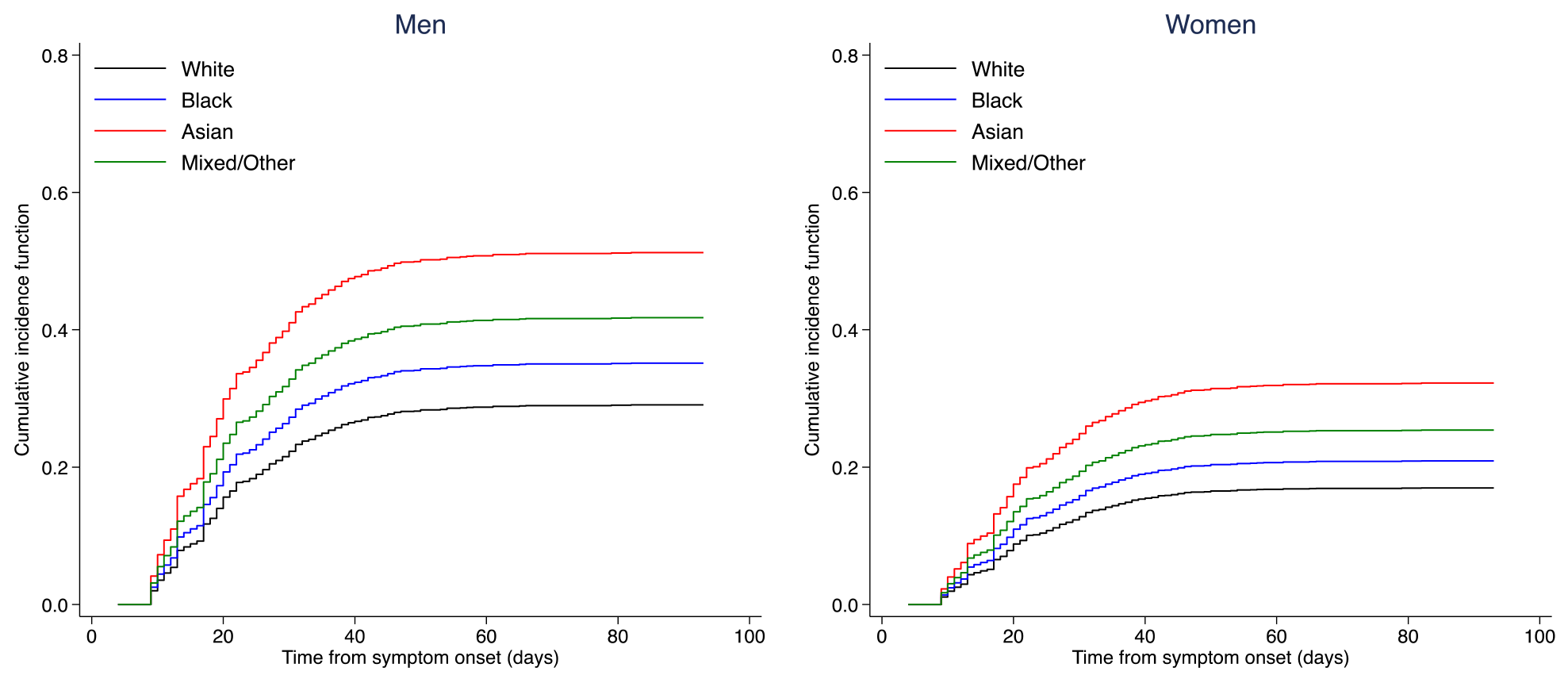
Cumulative age-adjusted incidence of ICU admission by sex and ethnicity

**Supplementary Figure 6.**
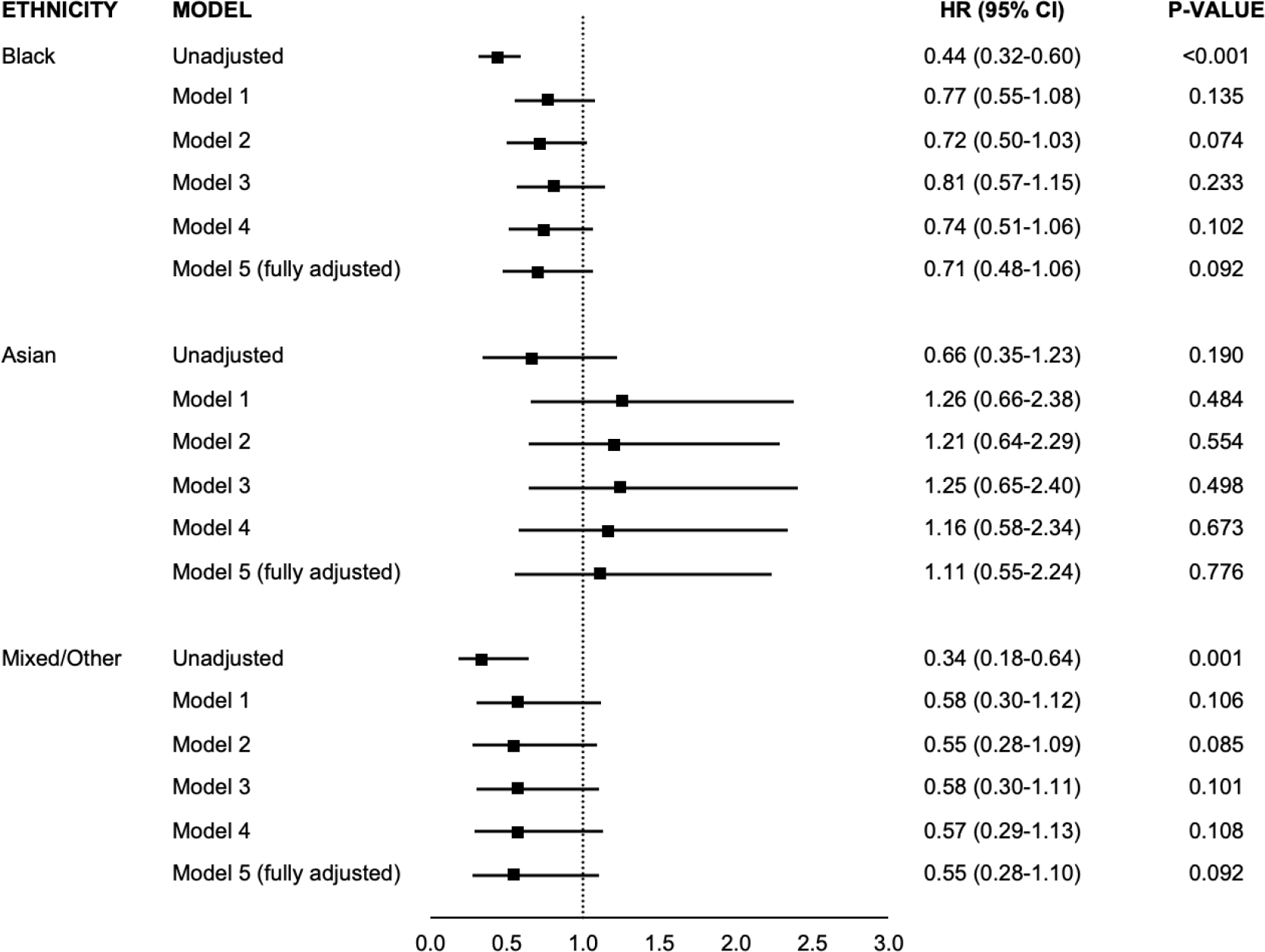
Association between ethnicity and risk of in-hospital mortality with COVID-19 in patients with BMI data available. Hazard ratios are compared to White ethnicity Model 1 – adjusted for age and sex Model 2 – adjusted for age, sex and index of multiple deprivation (IMD) Model 3 – adjusted for age, sex and body mass index (BMI) Model 4 – adjusted for age, sex and comorbidities* Model 5 (fully adjusted model) – adjusted for age, sex, IMD, BMI, and comorbidities *Comorbidities include asthma, chronic obstructive pulmonary disease, coronary heart disease, hypertension, diabetes, chronic kidney disease.

**Supplementary Figure 7.**
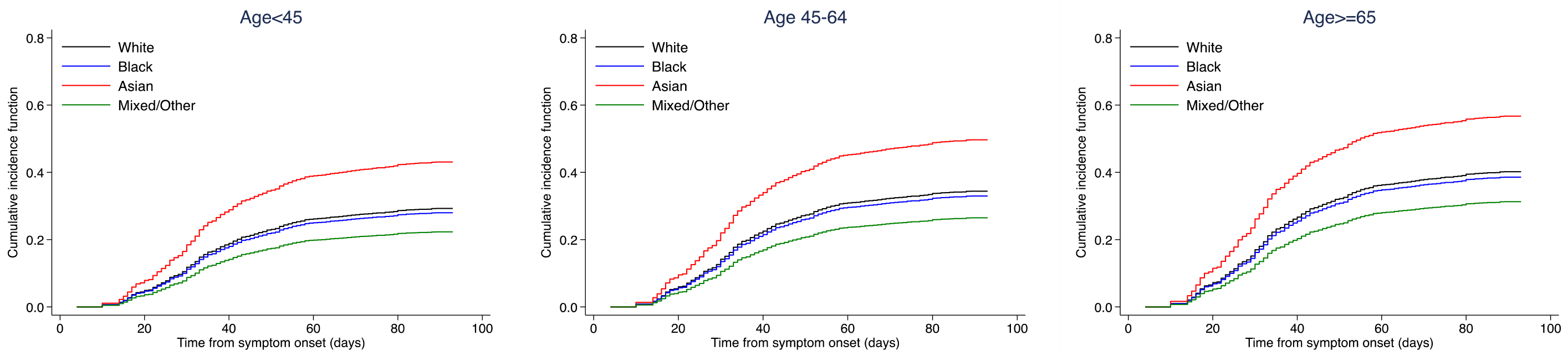
Cumulative age-adjusted incidence of in-hospital mortality by age-strata

